# Unexplained longitudinal variability in COVID-19 antibody status by Lateral Flow Immuno-Antibody testing

**DOI:** 10.1101/2021.08.23.21261638

**Authors:** Katrina Davis, Carolin Oetzmann, Ewan Carr, Grace Lavelle, Daniel Leightley, Michael Malim, Valentina Vitiello, Alice Wickersham, KCL-CHECK team, Reza Razavi, Matthew Hotopf

## Abstract

**Background:** COVID-19 antibody testing allows population studies to classify participants by previous SARS-CoV-2 infection status. Home lateral flow immune-antibody testing devices offer a very convenient way of doing this, but relatively little is known about how measurement and antibody variability will affect consistency in results over time. We examined consistency by looking at the outcome of two tests three months apart while COVID-19 infection rates were low (summer 2020 in the UK).

**Methods:** The KCL-Coronavirus Health and Experiences in Colleagues at King’s is an occupational cohort of staff and postgraduate research students. Lateral flow immune-antibody testing kits were sent to participant’s homes in late June 2020 and late September 2020. Participants also completed regular surveys that included asking about COVID-19 symptoms and whether they thought they had been infected.

**Results:** We studied 1489 participants returned valid results in both June and September (59% of those sent kits). Lateral flow immune-antibody test was positive for 7.2% in June and 5.9% in September, with 3.9% positive in both. Being more symptomatic or suspecting infection increased the probability of ever being positive. Of those positive in June, 46% (49/107) were negative in September (seroreversion), and this was similar regardless of symptom characteristics, suspicion, and timing of possible infection. A possible outlier was those aged over 55 years, where only 3 of 13 (23%) had seroreversion.

**Discussion:** These results do not follow the pattern reported from studies specifically designed to monitor seropositivity, which have found greater consistency over time and the influence of presence, timing and severity of symptoms on seroreversion. We suggest several factors that may have contributed to this difference: our low bar in defining initial seropositivity (single test); a non-quantitative test known to have relatively low sensitivity; participants carrying out testing. We would encourage other studies to use these real-world performance characteristics alongside those from laboratory studies to plan and analyse any antibody testing.

## Background

Antibody testing in the community has been used to indicate prior infection with SARS-CoV-2, the virus that causes COVID-19. This is important for monitoring and understanding the long-term consequences of SARS-CoV-2 infection. Seropositivity to SARS-CoV-2 may also identify individuals more likely to be immunologically protected in future waves of COVID-19. Lateral flow immuno-antibody (LFIA) testing for immunoglobulin M and G (IgM/IgG) at point-of-care or sent to people’s homes offers speed and convenience, with tests against gold-standard enzyme-linked immunoassay (ELISA) showing good accuracy of LFIA under lab conditions.(Pickering, Betancor et al. 2020) We previously reported the antibody status in June 2020 of participants in KCL-Coronavirus Health and Experiences in Colleagues at King’s – KCL-CHECK, a cohort of staff and postgraduate research students (PGRs) at a large UK university - using SureScreen LFIA cassettes detecting IgG/IgM antibodies to the SARS-CoV-2 spike protein (Ig-S).(Leightley, Vitiello et al. 2020, Davis, Carr et al. 2021). Comparison to other COVID-19 antibody studies suggested that the home use of LFIA in our cohort may have missed cases that would have been positive by more sensitive tests. Lower sensitivity may be an acceptable trade-off to avoid false positives and maximise usability.

One aspect of LFIA performance that remains unclear is consistency over time. To investigate this, we report on follow-up testing in the same cohort in September 2020 to look for inconsistencies that might be attributable to measurement and antibody variability. This will help to guide future users of home LFIA testing.

## Methods

Elsewhere described(Davis, Stevelink et al. 2020, Davis, Carr et al. 2021) KCL-CHECK is a cohort of staff and postgraduate research students at a large university in London, United Kingdom. SureScreen LFIA devices and equipment needed to perform the test were sent to participants’ home addresses in June 2020 and September 2020 (see Table 1). After completing the test, participants uploaded photographs of the results for interpretation by the research team, with any line in the control plus IgG and/or IgM being interpreted as positive (see supporting information Appendix S1 for the protocol and pictures of positive, negative and invalid test cassettes at Step 7). The only difference in the protocol between the two time-points was in how the buffer was supplied (see Appendix S1). Our previous paper compared the cohort with antibody results to the known characteristics of staff and PGRs, showing that women and people with White ethnicity were over-represented.(Davis, Carr et al. 2021)

**Table 1.** Timing of components of KCL-CHECK including two testing windows

| Date* | Time Period | Survey type for COVID-19 symptoms and suspicion | LFIA mailout |
| --- | --- | --- | --- |
| 16/04/2020 | T0 | baseline ("since the start of the pandemic") |  |
| 04/05/2020 | T1 | fortnightly ("in the last two weeks") |  |
| 18/05/2020 | T1 | fortnightly ("in the last two weeks") |  |
| 01/05/2020 | T1 | fortnightly ("in the last two weeks") |  |
| 15/06/2020 | T1 | 2 monthly ("in the last two months") | **22 – 30 Jun** |
| 29/06/2020 | T2 | fortnightly ("in the last two weeks") |  |
| 13/07/2020 | T2 | fortnightly ("in the last two weeks") |  |
| 27/07/2020 | T2 | fortnightly ("in the last two weeks") |  |
| 10/08/2020 | T2 | 2 monthly ("in the last two months") |  |
| 24/08/2020 | T2 | fortnightly ("in the last two weeks") |  |
| 07/09/2020 | T2 | fortnightly ("in the last two weeks") | **15 – 23 Sep** |

\* Date survey opened. Email alert sent to the majority on day one, and a smaller subset on day eight. Survey closed on day 13.

We collected general information about participants at baseline (April 2020). Core symptoms, symptom algorithm status and suspicion of COVID-19 were collected by survey. All participants in the antibody trial agreed to follow-up surveys every two months, with the majority also agreeing to surveys every two weeks. Any one of the following symptoms was considered a “core” symptom: a new persistent cough; fever; loss of smell or taste. More severely affected participants were identified by a symptom algorithm that also takes into accountsevere fatigue, loss of appetite, age and sex,(Menni, Valdes et al. 2020). Participants who thought they had “probably” or “definitely” experienced COVID-19 themselves were taken to indicate COVID-19 suspicion.

We present the survey results in three time periods: T0 February-April (pre-baseline), T1 May-June, and T2 June-September. The LFIA were sent out at the date indicated, but we accepted uploaded results for approximately a month after this, and some participants were mailed replacement kits at a later date. We have, however, used the first date possible, as this will ensure that, for example, exposure in T1 was very likely to have occurred before testing for antibodies in T1. Where participants have reported COVID-19 symptoms in multiple time-periods, we have taken the earlier report, to avoid persistent symptoms of COVID-19 being counted as a new infection. Participants were not excluded based on their completion of follow-up surveys. COVID-19 symptoms and suspicion were considered as ‘ever reported’ or ‘not reported’ at T0 and across all completed surveys in T1, and again in T2, with no completion in a survey being treated as ‘not reported’.

## Results

Most (2544, 91%) of the 2807 baseline cohort were eligible and elected to be sent an LFIA test. 1882 returned valid results for T1 and 1674 for T2, with 1489 (59% of sent, 53% of baseline) having valid results in both rounds of testing (see supporting information Figure S1), becoming our sample for analysis. The participant characteristics show that, compared to our baseline cohort, this sample under-represents PGRs and people with ethnicities other than White (see supporting information Table S1). The COVID-19 questions in each fortnightly survey were answered by between 92% to 82% of the sample (mean 88%). 58% (860/1489) of the sample answered at all ten surveys, 75% answered at least nine, and 92% answered at least five.

Table 2 shows that 107 of 1489 (7.2%) were positive at T1 and 83 (5.6%) were positive at T2, with only 58 (3.9%) positive at both. The number and timing of COVID-19 symptoms (core +/- algorithm) or suspicion are shown in supporting information Table S2. 715 participants (48%) reported symptoms and/or suspicion of COVID-19 at any time. Participants were much more likely to report COVID-19 symptoms or suspicion at T0 compared to T1 and T2, and most of those reporting suspicion or symptoms at T1 or T2 had already reported them at T0.

**Table 2.** Valid results from first two antibody testing windows in KCL-CHECK

|  |  | June 2020 (T1) LFIA result |  |  |
| --- | --- | --- | --- | --- |
|  |  | Positive | Negative | total |
| September (T2) LFIA result | Positive | 58 | 25 | **83** |
|  | Negative | 49 | 1357 | **1406** |
|  | **total** | **107** | **1382** | **1489** |

Table 3 and supporting information Figure S2 show the sequence of LFIA test results at T1 (positive/negative) and T2 (positive/negative) according to symptom algorithm, core symptoms and suspicion. The highest proportion of positive-positive occurred among those who were above cut-off on the symptom algorithm at T0 (17%). Inconsistent test results (either positive-negative or negative-positive) were common, at 56% of ever positive (74/132), and possibly more so for those presenting with COVID-19 symptoms or suspicion for the first time during T1 or T2. But only two of the 25 participants that converted from negative to positive had symptoms after T0, so COVID-19 infection shortly before the first test (T1) or between the first and second (T2) seems unlikely to account for this late seroconversion. There appears to be an increase in the proportion ever testing positive with increasing number of types of positive covid indicators. Of those who had not reported any symptoms or suspicion, 2.5% were ever seropositive, while of those with all three indicators (core symptoms, severe enough to score on the symptom algorithm and they suspected COVID-19 infection), 40% were ever seropositive. Of 107 with positive antibody results in T1, seropositivity was retained in T2 by 55% (58/107), which has a 95% confidence interval of 45-63%. Table 4 and supporting information Figure S3 show that the retention of antibodies for most subgroups was within the confidence interval of the sample. Outliers are the group aged 55+ years (above average at 77%). Those with a positive antibody result in T1 whose first symptoms or suspicion were reported closer to the first testing (in T1) do not seem more likely to retain seropositivity (in fact the opposite), but there are small numbers in those groups. Supporting information Figure S4 and Figure S5 illustrate that more COVID-19 indicators also do not seem to lead to greater retention of seropositivity, as asymptomatic (no symptoms or suspicion at T0 or T1) have a similar retention proportion (64%) to those with one indicator (63%) and to having multiple indicators (52%).

**Table 3.** Sequence of LFIA IgG/IgM results by COVID-19 indicators and timings (also graphed in Figure S2)

| Subgroup | First reported | number in subgroup | Proportion with sequence in group (T1-T2) |  |  |  |
| --- | --- | --- | --- | --- | --- | --- |
|  |  |  | Positive-Positive | Positive-Negative | Negative-Positive | Negative-Negative |
| **Overall** | Any | 1489 | 3.9% | 3.3% | 1.7% | 91.1% |
| Symptom algorithm | Ever | 256 | 14.1% | 14.1% | 3.5% | 68.4% |
|  | T0 | 206 | 17.0% | 15.5% | 3.9% | 63.6% |
|  | T1 | 30 | 3.3% | 10.0% | 3.3% | 83.3% |
|  | T2 | 20 | 0.0% | 5.0% | 0.0% | 95.0% |
| Core Symptoms* | Ever | 652 | 7.4% | 6.6% | 2.1% | 83.9% |
|  | T0 | 541 | 8.5% | 7.4% | 2.2% | 81.9% |
|  | T1 | 59 | 1.7% | 5.1% | 3.4% | 89.8% |
|  | T2 | 52 | 1.9% | 0.0% | 0.0% | 98.1% |
| Suspected COVID-19 | Ever | 409 | 11.7% | 10.0% | 3.4% | 74.8% |
|  | T0 | 357 | 12.9% | 10.1% | 3.9% | 73.1% |
|  | T1 | 40 | 0.0% | 7.5% | 0.0% | 92.5% |
|  | T2 | 12 | 16.7% | 16.7% | 0.0% | 66.7% |
| None of the above | Ever | 774 | 0.8% | 0.4% | 1.3% | 97.5% |
| Exactly 1 indicator | Ever | 298 | 1.7% | 1.3% | 0.7% | 96.3% |
| Exactly 2 indicators | Ever | 232 | 6.0% | 4.3% | 1.7% | 87.9% |
| All 3 indicators | Ever | 185 | 17.8% | 17.3% | 4.9% | 60.0% |

\* all participants with symptom algorithm are also in core symptoms

**Table 4.** Retention of seropositivity, as a proportion of participants testing positive at T1 who remained positive at T2 (from those with valid test results at both times, n=1489)

|  |  |  | Number in group | Number positive antibody T1 |  | Of which, number remain positive T2 (% of T1 @ T2) |
| --- | --- | --- | --- | --- | --- | --- |
| **Total** |  |  | 1489 | 107 | 7% | 58/107 54% |
| **Demographics** | Gender | Female | 1055 | 74 | 7% | 40/74 54% |
|  |  | Male | 424 | 31 | 7% | 17/31 55% |
|  |  | Other / NA | 10 | 2 | 20% | 1/2 50% |
|  | Age group (years at baseline) | 16-34y | 606 | 54 | 9% | 29/54 54% |
|  |  | 35-54y | 658 | 40 | 6% | 19/40 48% |
|  |  | 55+y | 225 | 13 | 6% | 10/13 77% |
| **COVID-19 symptoms** | Symptom algorithm | T0 | 206 | 67 | 33% | 35/67 52% |
|  |  | T1 | 30 | 4 | 13% | 1/4 25% |
|  | Core symptoms<sup>+</sup> | T0 | 541 | 86 | 16% | 46/86 53% |
|  |  | T1 | 59 | 4 | 7% | 1/4 25% |
|  | No core symptoms T0/T1 |  | 837 | 16 | 2% | 10/16 63% |
| **Suspect COVID-19** | Suspected covid-19 | T0 | 357 | 82 | 23% | 46/82 56% |
|  |  | T1 | 40 | 3 | 8% | 0/3 0% |
|  | Not suspected T0/T1 |  | 1080 | 18 | 2% | 10/18 56% |

*T0: first reported at baseline survey*
*T1: first reported in surveys after baseline and during T1*
*<sup>+</sup> All symptom algorithm positive participants are also in Core symptoms*

## Discussion

We report two rounds of antibody testing in an occupational cohort using LFIA at home after the first wave of COVID-19 in the UK. There was a variability in antibody test results obtained three months apart (timepoints T1 and T2) that was unexpected and not readily explained by the reported personal experience of COVID-19. 107 of 1489 (7.2%) participants tested positive at T1, with 58 (55%) remaining positive and 49 (45%) becoming negative at T2. Twenty-five participants who were negative at round one became newly positive in round two, accounting for 30% of those positive at T2, and not accounted for by COVID-19 symptoms or suspicion between testing time-points. Severity of symptoms of COVID-19 was related to the likelihood of ever testing positive, but not of retaining seropositivity. Recency of infection and gender did not affect proportion remaining positive, but there was a suggestion of older age groups being more likely to remain seropositive at T2.

The timing of symptoms and suspicion of COVID-19 in our sample accords with the known features of the early stages of epidemic in the UK,(Ward, Cooke et al. 2020) with most suspected infections in our population occurring before the end of April 2020. Our first LFIA testing directed towards anti-spike antibodies (primarily IgG-S) was in June. IgG-S levels are known to rise to a maximum level at around a month after infection.(Gudbjartsson, Norddahl et al. 2020, Korte, Buljan et al. 2020) Most studies have found levels declining after this,(Ibarrondo, Fulcher et al. 2020, Wu, Liu et al. 2020, Glück, Grobecker et al. 2021, Ward, Cooke et al. 2021) although others describe more of a plateau.(Gudbjartsson, Norddahl et al. 2020, Ortega, Ribes et al. 2021) In studies following IgG-S positive people for up to six months, some found little decline in seropositivity (García-Abellán, Padilla et al. 2021, Griffin, Tully et al. 2021), while some showed a substantial decline (Muecksch, Wise et al. 2020, Self 2020, Glück, Grobecker et al. 2021, Perez-Saez, Zaballa et al. 2021). In this analysis, among seropositive participants in June, 45% were negative in September. This exceeds levels of reversion seen in other studies, but it should be noted that these were usually restricted to people who had also tested positive for antigen or had repeated positive antibody tests. Compared to those with symptoms/suspicion in the first wave (T0, up to mid-April 2020), those few in our study who first reported symptoms/suspicion in T1 (April-June 2020) and tested positive in June were no more likely to remain seropositive in September, which is the opposite of what would be expected if recency of infection was playing a large part in the variability.

Use of the SureScreen LFIA and ELISA in hospitalised patients showed that IgG-S titres were more reliably detected in people with more severe COVID-19 illness.(Korte, Buljan et al. 2020, Wu, Liu et al. 2020) Other studies have shown that people who are symptomatic for COVID-19 remain seropositive more consistently than those who had an asymptomatic infection.(Ward, Cooke et al. 2020) We had three levels of COVID-19 symptoms among our sample: no core symptoms reported, core symptoms and algorithm positive. There was no evidence of increases in retention of seropositivity in more symptomatic cases, as retention was 63% with no core symptoms 52% with core symptoms and 52% in those with a positive algorithm. IgG-S titres have also previously been found to be higher in men than women and in older age groups,(Korte, Buljan et al. 2020, Wu, Liu et al. 2020) although they may subsequently fall faster.(Ward, Cooke et al. 2021) In our study, older participants were more likely to maintain seropositivity, which could be the outcome of having higher antibody titres after exposure but could be an artefact due to low numbers.

There is known to be substantial heterogeneity of COVID-19 antibody test results depending on the type of antibody probed and the kit used.(Kontou, Braliou et al. 2020, Favresse, Eucher et al. 2021, Jones, Mulchandani et al. 2021). For some LFIAs, heterogeneity has additionally been shown between batches of testing kits, lab versus point of care use, and the use of venous versus capillary blood.(Flower, Brown et al. 2020, Silveira, Mesenburg et al. 2021) Although the LFIA result have a very predictable relationship to antibody titres under tightly controlled conditions,(Ward, Cooke et al. 2021) in practice minor variations in the use of the LFIA, especially for people with modest levels of immunoglobulin, may give results that are above and below cut-off at the same actual immunoglobulin titre.(Favresse, Eucher et al. 2021, Silveira, Mesenburg et al. 2021) This may have contributed to our unexpected findings.

The strengths of this study are the use of a longitudinal cohort, allowing comparison of sequential testing, and COVID-19 indicators dating to the first wave of the pandemic, with good completion of frequent follow-up questionnaires. The same antibody test, batch of tests and near-identical protocol was used for both testing episodes, reducing variability from these causes. The mailing of kits allowed for greater participation at a time of heightened health concerns, but may have increased variability of use for a test kit that was designed for use by a healthcare professional. Despite the size of the cohort, we found numbers testing positive were low, which has affected the statistical power we could achieve. We were not able to look at antibody results in people severely affected by COVID-19 (i.e. requiring hospitalisation) as there were very low numbers. We did not perform antigen testing at the time of symptoms nor laboratory tests of antibodies. Participants were able to report tests done externally, but that data has not been shown here as access to testing was inconsistent and may bias results.

## Conclusion

There is variability in people identified as seropositive through LFIA home testing over time that is not easily explained by symptom severity or timing. Therefore, we suggest that home antibody testing should be considered in the context of self-reported measures in research and the full presentation in clinical settings.

## Data Availability

Researchers may apply to have access to pseudonymised data. Requests to access study data is subject to submission of a research proposal to the Principal Investigators (Professor Matthew Hotopf, Professor Reza Razavi and Dr Sharon Stevelink). All requests must be made in accordance with the UK Policy Framework for Health and Social Care research. Where the applicant is outside of Kings College London a data-sharing agreement is required.

