## Supplementary Materials for "Unexplained longitudinal variability in COVID-19 antibody status by Lateral Flow Immuno-Antibody testing"

**Part 1: Extra Figures and Tables**

Figure S1 Participant Flowchart

Figure S2 Bubble chart of the relationship of probability seropositive at T1 versus remaining seropositive at T2, according to (1) symptoms or suspicion status, orange; (2) gender, yellow; (3) age, green. Size of bubble reflects number of people in subgroup

Figure S3 Profile of COVID indicators reported in T0 to T1 in those who were positive for antibodies at T1 and those positive at both T1 and T2

Figure S4 Numbers and proportion of those reporting 0, 1, 2 or 3 COVID indicators who were positive at two time-points

Table S1 Participation from baseline to sample for this follow-up antibody study

Table S2 Pattern of recording of three COVID-19 indicators over time-points

**Part 2: Appendix S1 – Procedure for LFIA testing**

Participant instruction sheet (including an explanation of valid and invalid tests, step 7)

Difference between procedures for testing at T1 and testing at T2

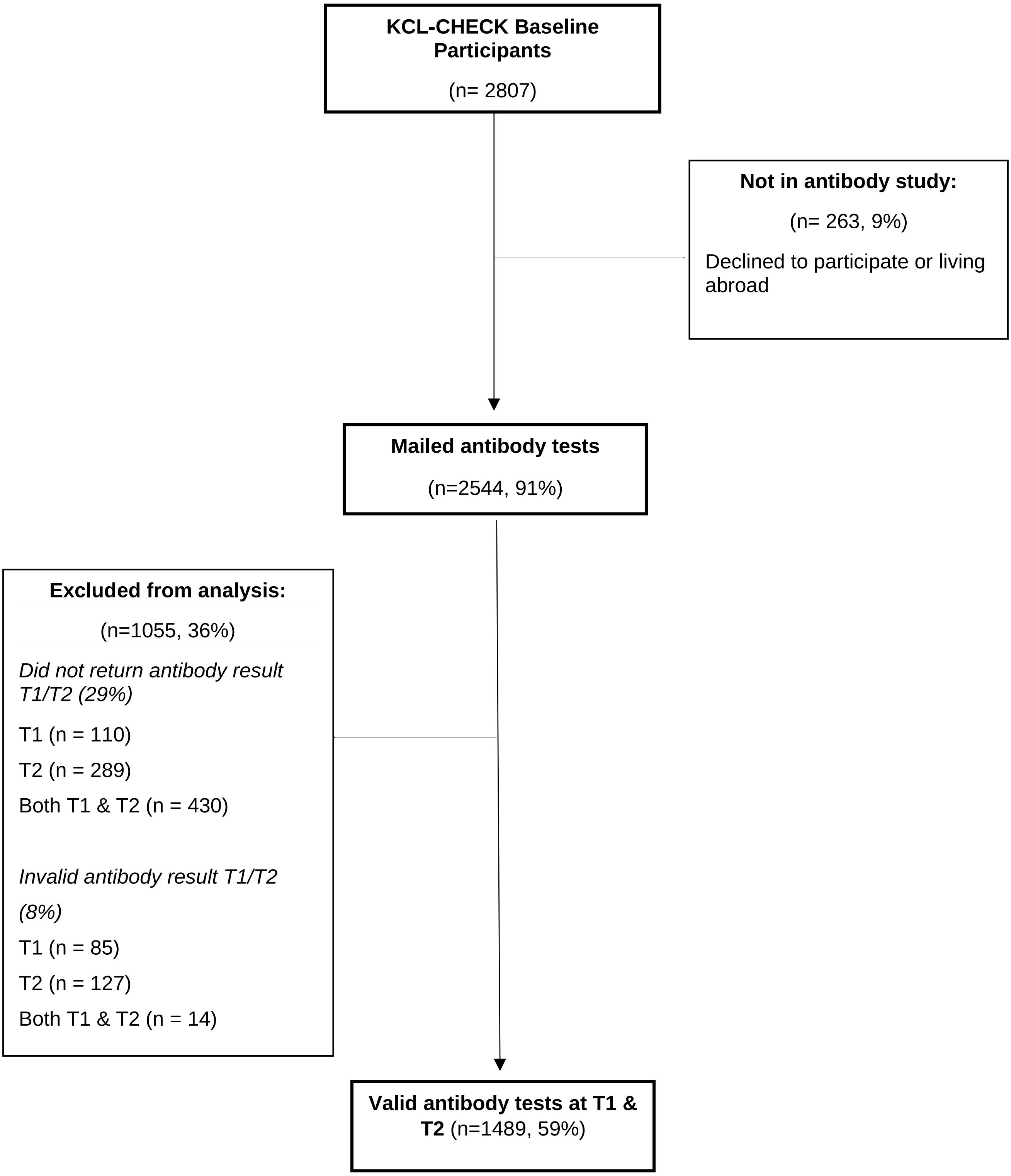


*Figure S1 Participant Flowchart*

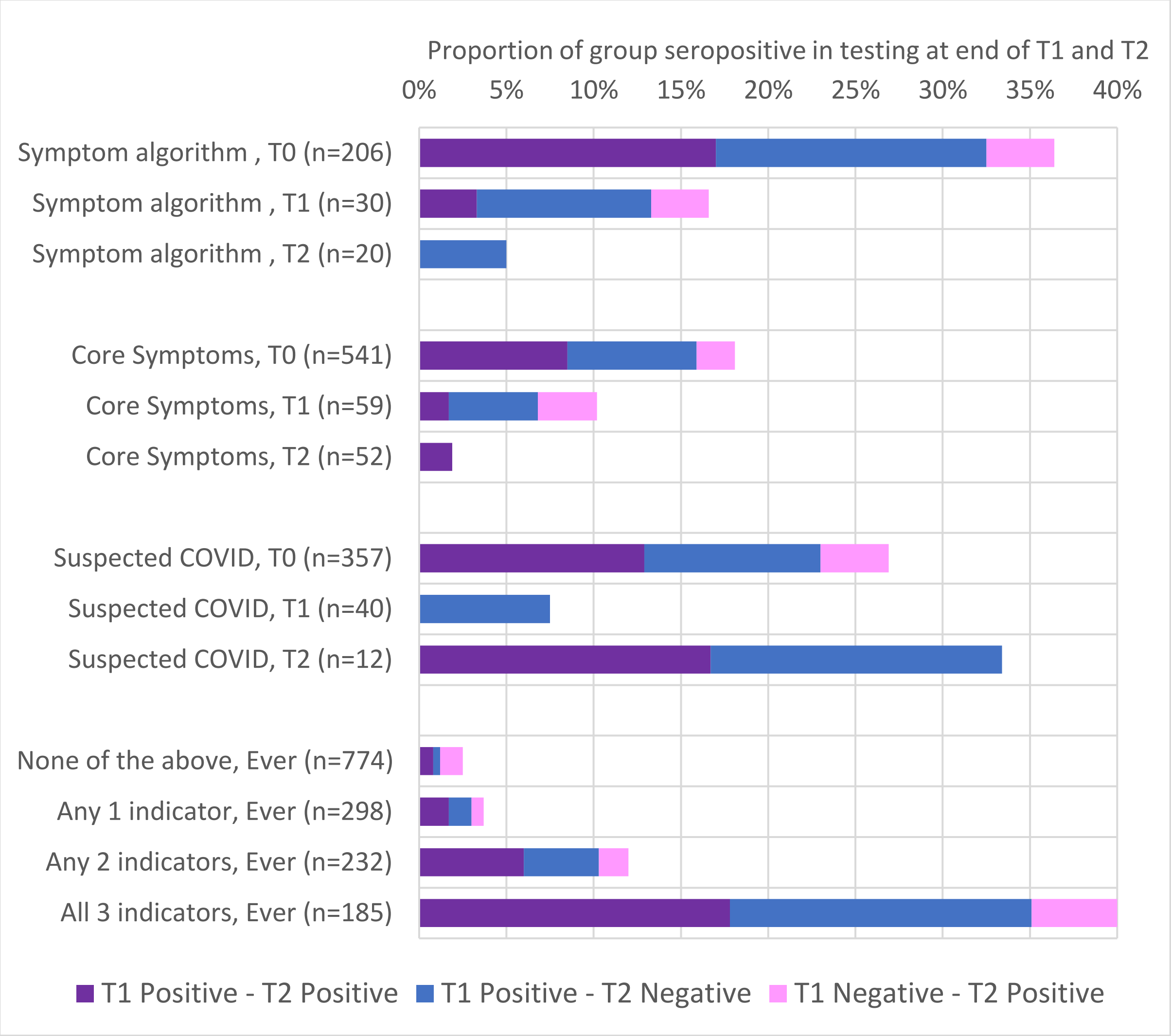


*Figure S2 Proportion of group seropositive and sequence of tests by COVID-19 indicators and timing (see table 3 for numbers)*

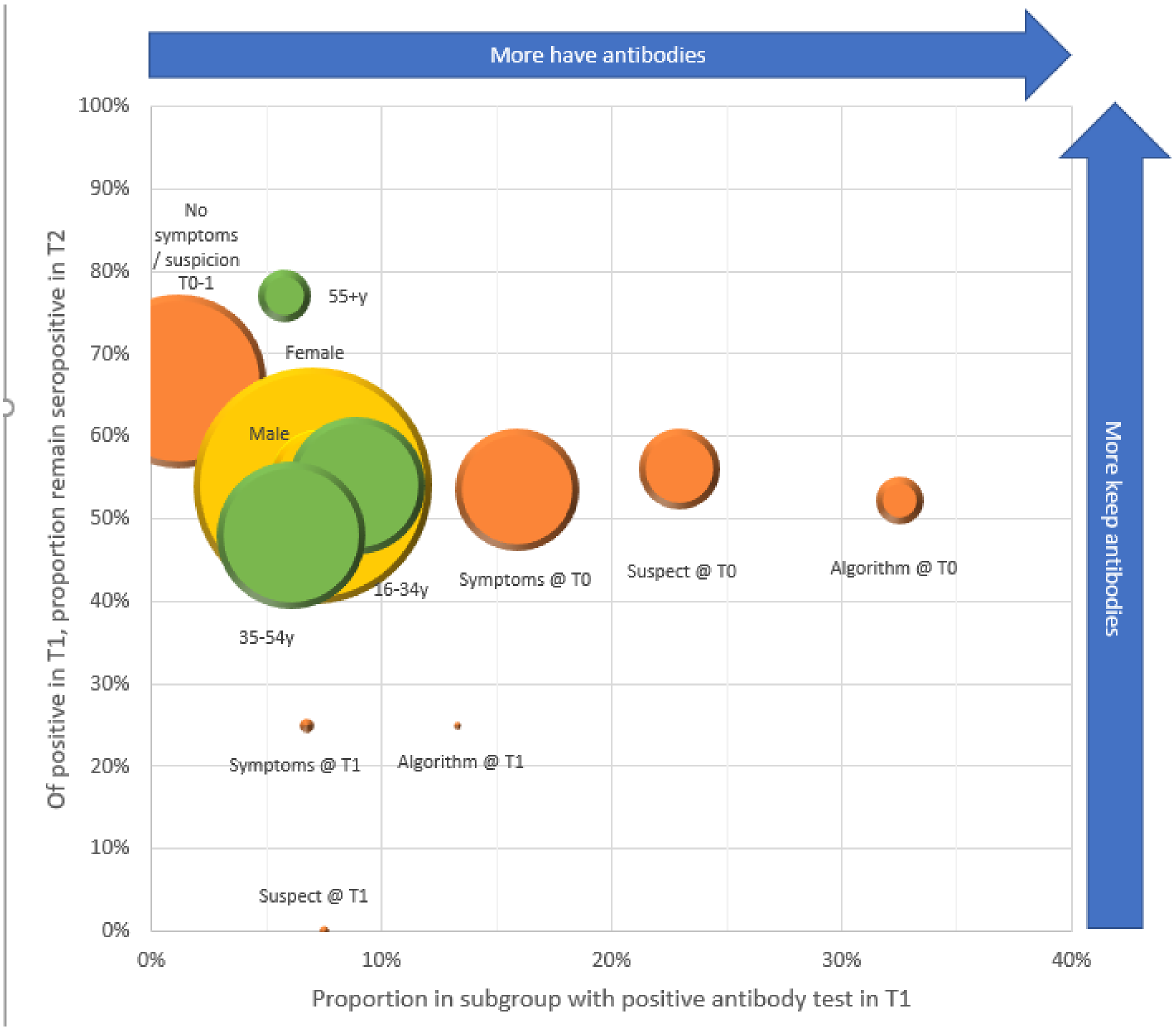


*Figure S3 Bubble chart of the relationship of probability seropositive at T1 versus remaining seropositive at T2, according to (1) symptoms or suspicion status, orange; (2) gender, yellow; (3) age, green. Size of bubble reflects number of people in subgroup.*

Footnotes: Algorithm = COVID-19 symptom algorithm positive; Suspect = reported probably or definitely had COVID-19; Symptoms = reported at least one of the core COVID-19 symptoms; T0 = reported at baseline; T1 = did not report at baseline, but reported before testing at T1

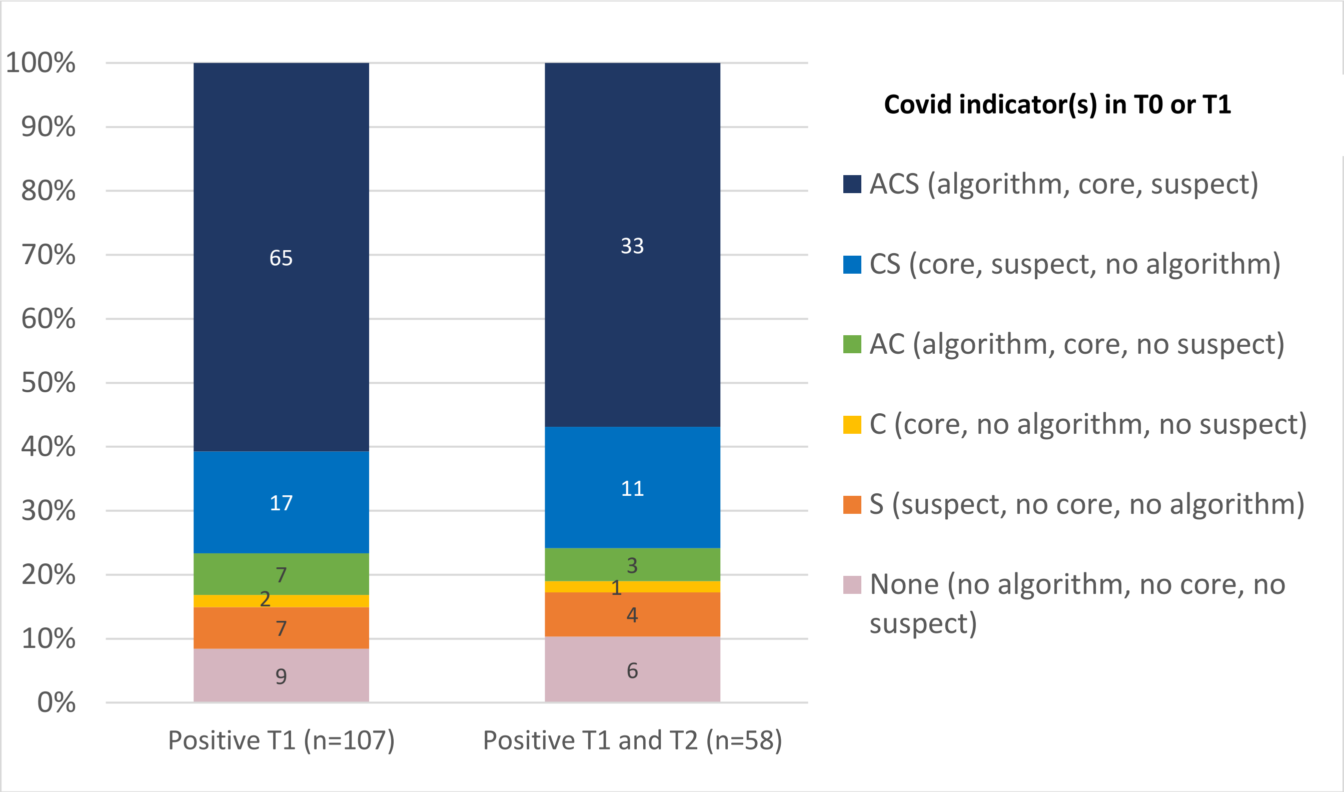


*Figure S4 Profile of COVID indicators reported in T0 to T1 in those who were positive for antibodies at T1 and those positive at both T1 and T2*

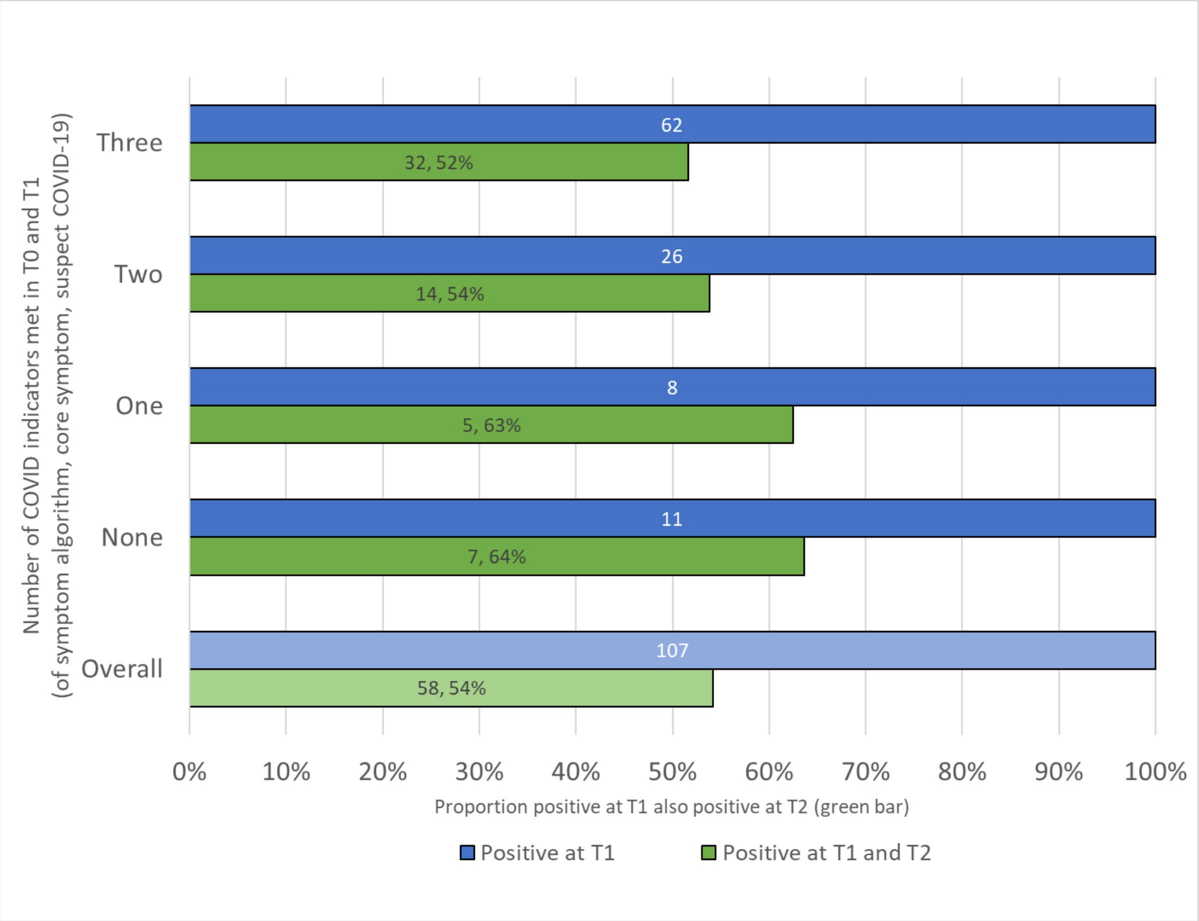


*Figure S5 Numbers and proportion of those reporting 0, 1, 2 or 3 COVID indicators who were positive at two time-points*

*Table S1 Participation from baseline to sample for this follow-up antibody study, with red shading for over-represented and blue shading for under-represented as compared to baseline*

| | | Baseline responses | LFIA testing cohort | Valid antibody result (June) | Two valid antibody results | Two results as proportion of baseline group |
|---|---|---|---|---|---|---|
| **Overall** | | **2807** | **2544** | **1882** | **1489** | **53%** |
| Gender | Female | 1948 (69%) | 1769 (70%) | 1339 (71%) | 1055 (71%) | 54% |
| | Male | 842 (30%) | 760 (30%) | 533 (28%) | 424 (28%) | 50% |
| Role | Academic, etc. | 1075 (38%) | 1060 (42%) | 810 (43%) | 643 (43%) | 60% |
| | Clerical etc. | 809 (29%) | 797 (31%) | 622 (33%) | 501 (34%) | 62% |
| | Clinical etc. | 204 (7%) | 199 (8%) | 134 (7%) | 108 (7%) | 53% |
| | PGRs | 536 (19%) | 452 (18%) | 315 (17%) | 236 (16%) | 44% |
| | missing | 183 (7%) | 36 (1%) | 1 (0%) | 0 (0%) | 0% |
| Age-group | 18-24 | 136 (5%) | 125 (5%) | 90 (5%) | 69 (5%) | 51% |
| | 25-34 | 1084 (39%) | 948 (37%) | 695 (37%) | 537 (36%) | 50% |
| | 35-44 | 762 (27%) | 698 (27%) | 535 (28%) | 425 (29%) | 56% |
| | 45-54 | 440 (16%) | 409 (16%) | 287 (15%) | 233 (16%) | 53% |
| | 55-64 | 307 (11%) | 288 (11%) | 220 (12%) | 181 (12%) | 59% |
| | 65+ | 78 (3%) | 76 (3%) | 55 (3%) | 44 (3%) | 56% |
| Ethnicity group | White | 2152 (77%) | 2178 (86%) | 1660 (88%) | 1323 (89%) | 61% |
| | Mixed | 102 (4.2%) | 46 (1.8%) | 29 (1.5%) | 19 (1.3%) | 19% |
| | Asian | 204 (7.3%) | 191 (7.5%) | 125 (6.6%) | 96 (6.4%) | 47% |
| | Black | 39 (1.4%) | 37 (1.5%) | 20 (1.1%) | 17 (1.1%) | 44% |
| | Other | 70 (2.5%) | 69 (2.7%) | 44 (2.3%) | 31 (2.1%) | 44% |
| | missing | 240 (8.6%) | 23 (1.0%) | 4 (0.2%) | 3 (0.2%) | 1% |
| Keyworker status | keyworker | 358 (13%) | 357 (14%) | 242 (13%) | 222 (15%) | 62% |
| | non-key | 2449 (87%) | 2187 (86%) | 1640 (87%) | 1267 (85%) | 52% |

*Academic etc.: Staff who report being in Academic, Management or Specialist role*

*Clerical etc.: Clerical, Research or Technical roles*

*Clinical etc.: Teaching, Clinical or Facilities role*

*PGR: Post-graduate research student*

*Table S2 Pattern of recording of three COVID-19 indicators over time-points*

| | | T0 | T1 | T2 | Never |
|---|---|---|---|---|---|
| **Symptom Algorithm Positive** | Criteria met **in time period** | 206 | 94 | 65 | 1233 |
| | Criteria met **for the first time** | 206 | 30 | 20 | 1233 |
| | also reported at earlier time | na | 64/94, 68% | 45/65, 69% | |
| **Core symptoms** | Reported **in time period** | 541 | 219 | 204 | 837 |
| | Reported **for the first time** | 541 | 59 | 52 | 837 |
| | also reported at earlier time | na | 160/219, 73% | 152/204, 75% | |
| **Suspect COVID** | Reported **in time period** | 357 | 157 | 39 | 1080 |
| | Reported **for the first time** | 357 | 40 | 12 | 1080 |
| | also reported at earlier time | na | 117/157, 75% | 27/39, 69% | |

*T0: At baseline (Feb-Apr), T1: After baseline but before antibody testing in June (Apr-Jun), T2: Reported around the time of or after first antibody testing and before the second test in September (Jun-Sept)*

### Appendix S1

#### Participant Protocols for LFIA use

Please see the following pages for the full protocol for the June mail-out.

Step 7 shows guide to valid and invalid results

#### Changes to the protocol in September

In June, buffer was provided by SureScreen in bulk, which KCL-CHECK staff split into a bottle for each participant, which the participant needed to pipette from the bottle to the test cassette.

In September, greater convenience was achieved by sourcing individual doses of the same buffer (of a separate batch) from SureScreen that participants could squeeze directly onto the test cassette. Step 5 was therefore altered as below:

**Step 5: Add the buffer to the test cassette**

Open the buffer tube and add the buffer onto the test cassette area **marked 'B'** by squeezing
You must **add two-three drops of buffer**, try to do it as quickly as possible.

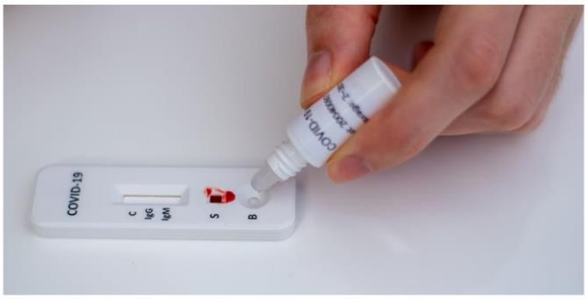

Dear [FirstName],

Thank you for your participation in the KCL CHECK (Coronavirus: Health & Experiences of Colleagues at King's) study. You are contributing to research concerning an unprecedented situation and are helping to improve understanding of how the COVID-19 (Coronavirus) pandemic is affecting the work, health and wellbeing of staff and postgraduate research students at King's College London.

As part of KCL CHECK, you consented to taking part in the virology component of the study and we are pleased to enclose your antibody home-testing kit. This test is comprised of a simple device used to detect the presence or absence of two types of COVID-19 antibodies (IgG and IgM) in a blood sample. Studies have shown the body produces these antibodies after experiencing COVID-19 infection. However, this is a research study and not a clinical test, so you should not rely only on the result of this test as either confirming that you have not had COVID-19 if the test is negative, or that you have had COVID-19 if the test is positive.

The test involves pricking your fingertip and taking a small blood sample which will be analysed in the test cassette supplied. This process should take around 10 minutes. You will need to photograph the result within 20 minutes of beginning the test and upload the picture to the KCL CHECK website: www.kcl-check.org/test-results.

Enclosed you will find:

1. A Coronavirus Rapid Test Cassette and kit (contents explained furtherin the instructions);
2. Step-by-step instructions for how to perform the test, how to interpret the result, andhow to share the result with the KCL CHECK research team. **Please** read these before conducting the test and follow the steps carefully.

You can find more information about the KCL CHECK study on the Participant Information Sheet at www.kcl-check.org/pis. The FAQ section of our website can also be found here: www.kcl-check.org/faq. If you have any further questions or if you would like to speak to the KCL CHECK Research Team, please.

Thank you for participating in this important research.

Best wishes,

KCL CHECK Research Team

**Please turn over for the step-by-step instructions when you are ready to begin the test**

**KCL CHECK**

**Instructions for COVID-19 antibody home-testing kit**

**(COVID-19 IgG/IgM Rapid Test Cassette)**

**Test kit contents:**

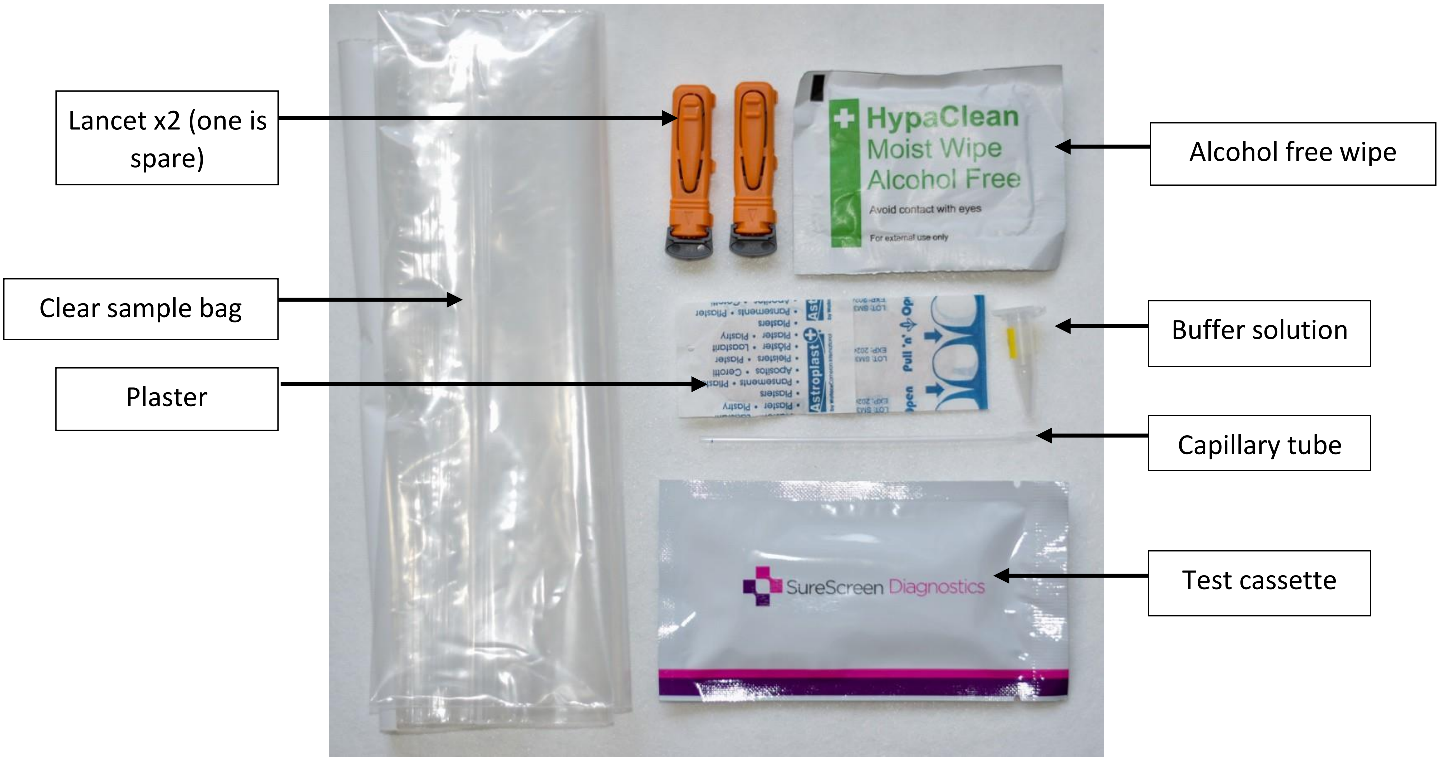


You also need a **<u>clean tissue</u>**, a **<u>timer</u>** and a way **<u>to photograph</u>** your result.

#### Step by step instructions

The test must be interpreted and photographed within <u>10-20</u> minutes of completingthe procedure to work correctly, so make sure you have this time before starting.

**Step 1: Set-up**

Before you begin, check you have everything you need (use the contents image above for reference). Then, remove the test cassette from the packaging **(use within 1 hour of opening)** and lay out the contents of your test kit.

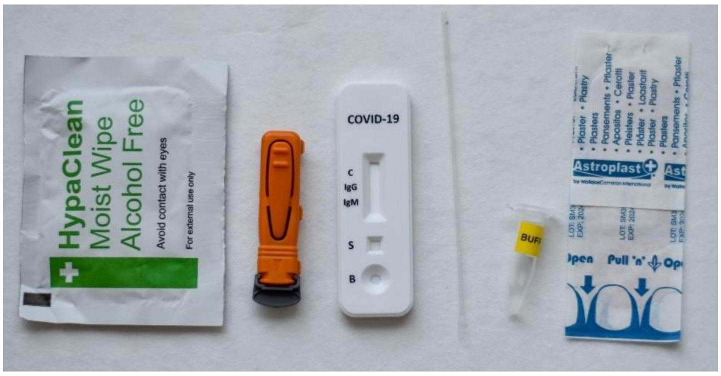

#### Step 2: Wash your hands and wipe your fingertip

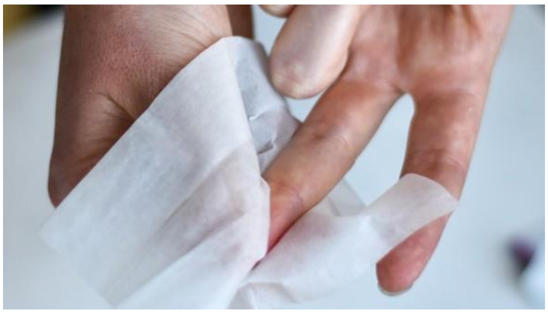

Using warm water, wash your hands. This helps with blood flow. Then, select either your middle or ring finger, and use the alcohol-free wipe to clean it.

#### Step 3: Puncture the skin using a lancet

To use the lancet, **twist** the grey cap clockwise, but **do not pull it**. As you twist, the cap will release on its own. Use the second lancet if you have problems with the first.

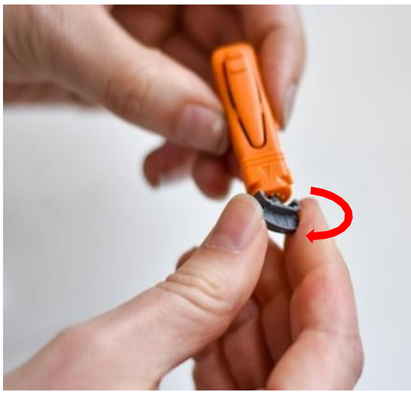

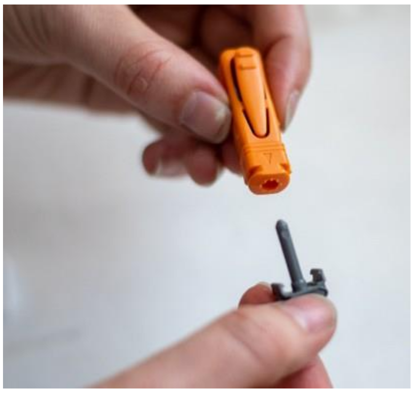

With the cap removed, massage your hand without touching the puncture site by rubbing down the hand towards the fingertip to encourage blood flow.

Then, hold the lancet firmly on your fingertip, and press the button to release the needle.

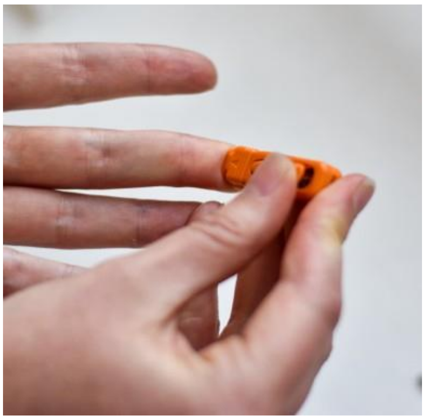

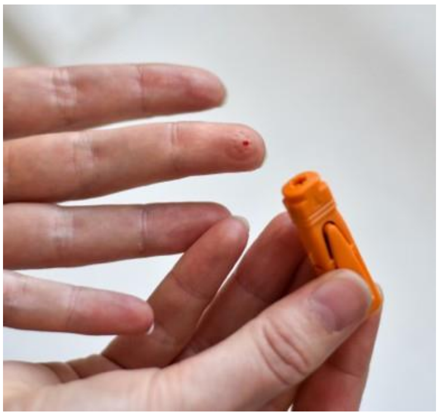

Wipe away the first sign of blood with a clean tissue.

#### Step 4: Add a drop of blood to the test cassette

Gently run your hand from the wrist up to the finger to form a rounded drop of blood. Release one drop of blood into the specimen well of the test cassette, **<u>this is marked ’S'</u>**.

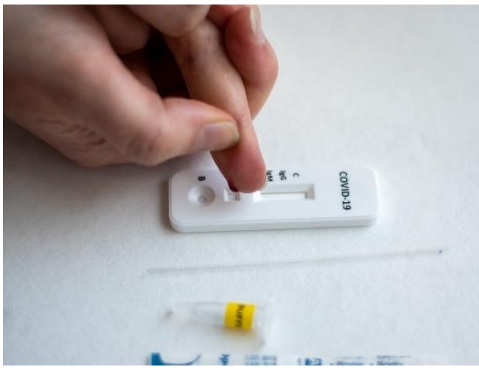

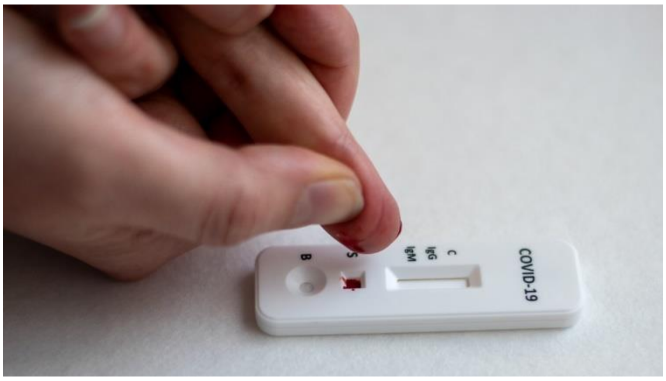


#### Step 5: Add the buffer to the test cassette

Open the buffer tube and add the buffer to the cassette using the capillary tube. To use the capillary, press firmly half-way down, then place the open end into the buffer and release. This will draw up the liquid into the capillary tube.

Then, drop the buffer solution onto the test cassette area **<u>marked ‘B’</u>** by squeezing the capillary tube. You must <u>use all the buffer</u> in the buffer tube, so you may have to repeat this process, try to do it as quickly as possible.

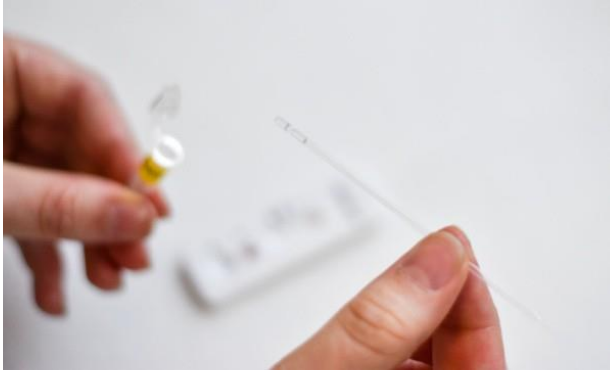

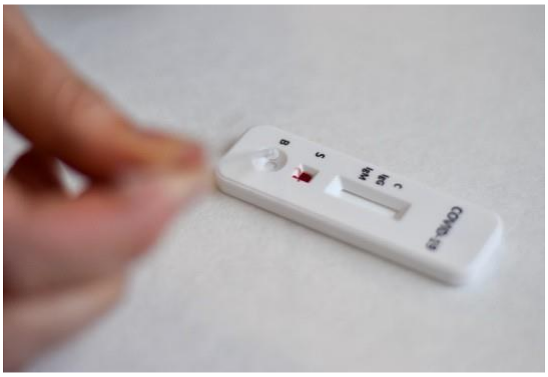

#### Step 6: Start the timer and record the current time

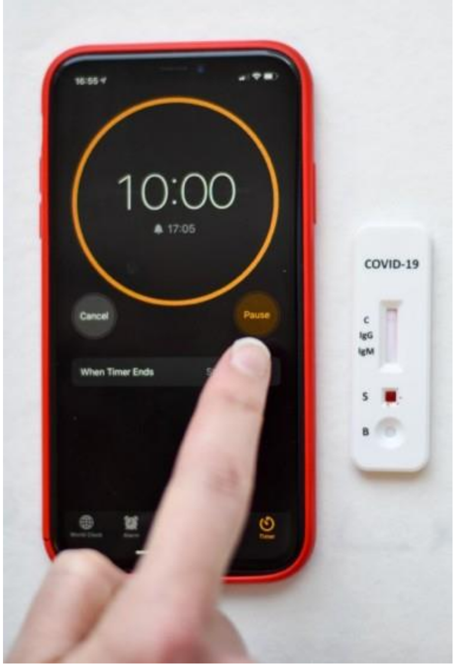


As soon as you have added the buffer, start a timer for **10 minutes**. Check the current time and record it in the box on the next page as the ‘test start time’. Please record the time in a 24-hour format (e.g.16:55).

While you wait, use the plaster to seal the puncture site on your fingertip.

#### Step 7: Check your result

After 10 minutes, coloured line(s) should appear on the test cassette. You can use the guide below to check your results.

**Result Interpretation Guide:** (See Figure 1)

**Note: The COVID-19 IgG/IgM Rapid Test Cassette will only indicate the presence of COVID-19 antibodies and should not be used as the <u>sole</u> criteria for the diagnosis of COVID-19. A negative result does not at <u>any time</u> exclude the possibility of COVID-19 infection. Similarly, a positive result <u>does not</u> confirm a previous COVID-19 infection. Please follow current NHS guidelines regarding COVID-19 symptoms and diagnosis.**

**IgG and IgM POSITIVE:** Three lines appear. A colored line appears each in the control region (C), the IgG test region and IgM test region. The result is positive for IgG & IgM antibodies and may indicate an immune response to a COVID-19 infection.

**IgG POSITIVE:** Two lines appear. A colored line appears in the control region (C), and also in the IgG test region. The result may indicate the presence of the specific antibody produced by the immune system in response to a COVID-19 infection.

**IgM POSITIVE:** Two lines appear. A colored line appears in the control region (C), and also in the IgM test region. The result may indicate the presence of the general antibody produced by the immune system as a first response to a COVID-19 infection.

**NEGATIVE**: One colored line appears in the control region (C). No lines appear in the IgG and IgM test regions. This may indicate no immune response present for a COVID-19 infection

**INVALID**: Control line fails to appear. This could happen for multiple reasons. Please ensure that you still record and share the results so that the research team are aware.

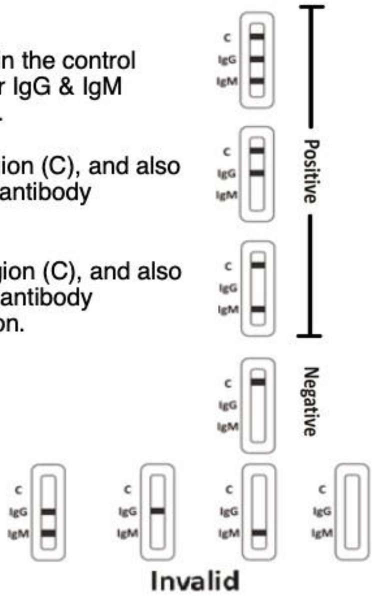


**Figure 1:** Result Interpretation

#### Step 8: Record your result

Without delay, place your test cassette in the box below and record your result, as well as the current time in ‘test end time’ (this should be in a 24-hour format).

| Place<br><br>test<br><br>cassette<br><br>here | Please complete the following based on your interpretation of the result:<br><br>☐<br>Positive ☐<br>Negative ☐<br>Invalid<br><br>Test start time:______:_____<br><br>Test end time:______:_____ |
|---|---|

Take a photograph of the result. Make sure that the whole box is visible, and that the imageis clear (see examples below). This must be done **within 20 minutes** of you completing thetest to generate a valid result – **do not check or photograph the result after 20 minutes.**

✓ The whole box is clearly visible, with all information complete.

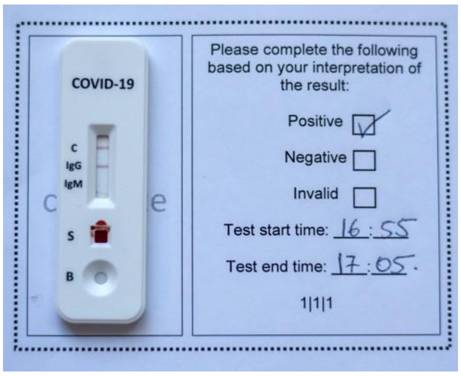


✗ Test obstructing information.

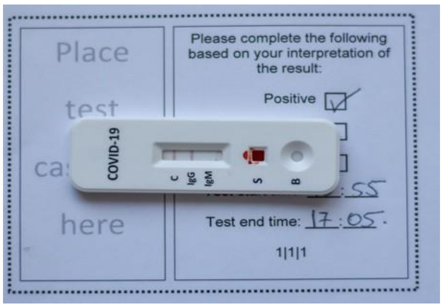


Whole box not visible.

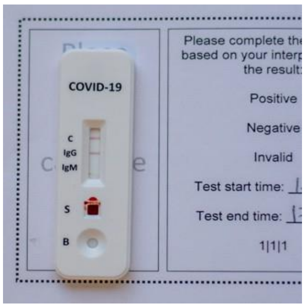


Image is blurry.

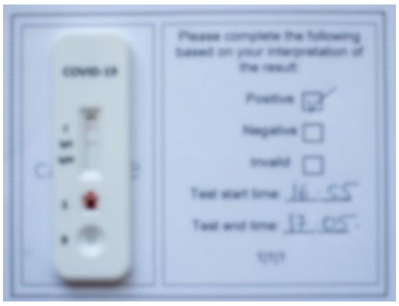

#### Step 9: Upload your photograph

Visit **kcl-check.org/test-results** via your web browser and upload and submit yourphotograph to share the test result with the KCL CHECK research team.

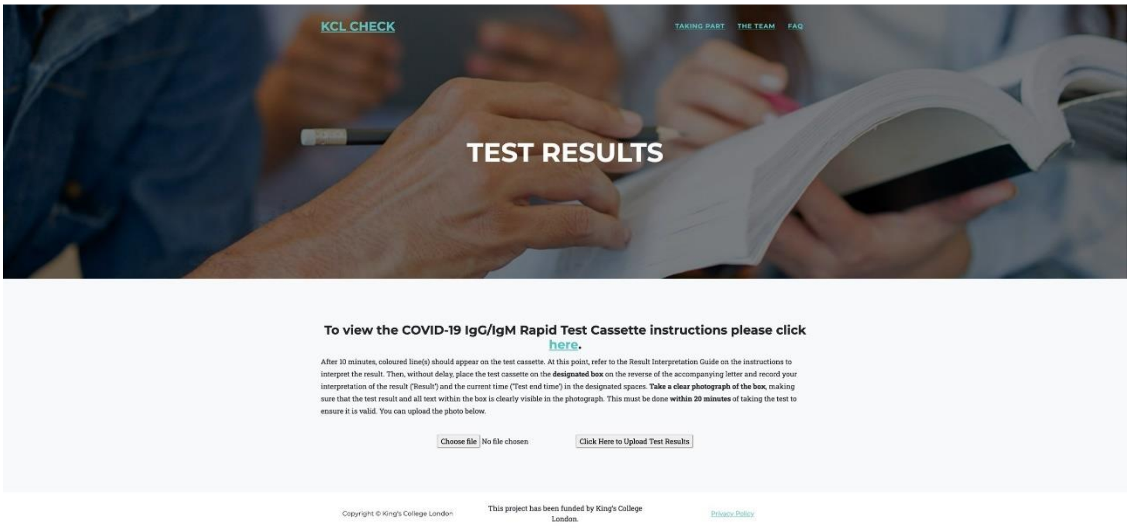


#### Step 10: Dispose of the kit in the clear sample bag

Place the used test cassette, buffer, lancet, capillary tube and alcohol wipe into the clearsample bag. **Seal the bag and dispose of in general waste.**
